# Ethnicity and outcomes in patients hospitalised with COVID-19 infection in East London: an observational cohort study

**DOI:** 10.1101/2020.06.10.20127621

**Authors:** V. J. Apea, Y. I. Wan, R. Dhairyawan, Z. A. Puthucheary, R. M. Pearse, C. M. Orkin, J. R. Prowle

## Abstract

**Background:** Preliminary studies suggest that people from Black, Asian and Minority Ethnic (BAME) backgrounds experience higher mortality from COVID-19 but the underlying reasons remain unclear.

**Methods:** Prospective analysis of registry data describing patients admitted to five acute NHS Hospitals in east London, UK for COVID-19. Emergency hospital admissions with confirmed SARS-CoV-2 aged 16 years or over were included. Data, including ethnicity, social deprivation, frailty, patient care and detailed risk factors for mortality, were extracted from hospital electronic records. Multivariable survival analysis was used to assess associations between ethnic group and mortality accounting for the effects of age, sex and various other risk factors. Results are presented as hazard ratios (HR) or odds ratios (OR) with 95% confidence intervals.

**Findings:** 1996 adult patients were admitted between 1^st^ March and 13^th^ May 2020. After excluding 259 patients with missing ethnicity data, 1737 were included in our analysis of whom 511 had died by day 30 (29%). 538 (31%) were from Asian, 340 (20%) Black and 707 (40%) white backgrounds. Compared to White patients, those from BAME backgrounds were younger, with differing co-morbidity profiles and less frailty. Asian and Black patients were more likely to be admitted to intensive care and to receive invasive ventilation (OR 1·54, [1·06-2·23]; p=0·023 and 1·80 [1·20-2·71]; p=0·005, respectively). After adjustment for age and sex, patients from Asian (HR 1·49 [1·19-1·86]; p<0·001) and Black (HR 1·30 [1·02-1·65]; p=0·036) backgrounds were more likely to die. These findings persisted across a range of risk-factor adjusted analyses.

**Interpretation:** Patients from Asian and Black backgrounds are more likely to die from COVID-19 infection despite controlling for all previously identified confounders. Higher rates of invasive ventilation in intensive care indicate greater acute disease severity. Our analyses suggest that patients of Asian and Black backgrounds suffered disproportionate rates of premature death from COVID-19.

**Funding:** None

**Research in context:** *Evidence before this study:* We searched PubMed, Google Scholar, Medrxiv, Trip Medical Database and internet search engines from inception to May 10^th^ 2020, using the terms “(COVID-19 or 2019-nCoV or SARS-CoV-2) AND (ethnicity)”, with no language restrictions, for research articles, editorials and commentaries. We identified 25 articles. Ten were international opinion pieces, fifteen were research articles reporting analyses of national and cohort datasets, predominantly in the United Kingdom (UK) and United States (US). Each of these studies indicated an increased risk of adverse outcomes in people from BAME backgrounds; either in terms of COVID-19 acquisition, disease severity or mortality. However, the underlying causes were unclear. Aggregated US data determined the relative risk of death for those of Black ethnicity compared to White ethnic groups to be 3.57. Three UK biobank cohort studies, limited by low BAME representation, described ethnicity as an independent risk factor of COVID-19 infection, partially attenuated by socio-economic status (SES). Analysis of a London hospital cohort of 520; experiencing 144 deaths, revealed an age and co-morbidity adjusted mortality odds ratio of 1.72 in Black populations of borderline significance. Age and geographical region-adjusted standardised mortality ratios, derived from UK composite hospital data, emphasised ethnic differences; being 2.41 for Bangladeshis and 3.24 for Black Africans. The impact of gender and deprivation was not explored. Another study of 5683 in-hospital deaths (England alone; 629 (11%) BAME) confirmed increased mortality risks in people from Black and Asian groups only partially attributable to social deprivation and co-morbidity but did not adjust for other vulnerability factors. There remained a need for a more detailed analysis of outcomes across different ethnic groups in a large, high acuity dataset, adjusting for broader clinical and laboratory prognostic factors, alongside SES, smoking status, age, body mass index (BMI) and sex.

*Added value of this study:* We conducted a large observational cohort study of COVID-19 hospital admissions within an area which experienced the highest rates of COVID-19 infection and mortality in the UK. It offers detailed insight into a majority (60%) ethnically diverse cohort and adds substantial evidence that ethnicity is a predictor of poor outcomes for COVID-19 patients at, and beyond, 30 days. Using robust multivariable survival analyses we have quantified and described the impact on this association of a number of additional prognostic factors such as frailty score and markers of inflammation alongside age, sex, deprivation, co-morbidity, BMI and smoking status. Those of Asian and Black ethnicities were consistently found to have an increased risk of 30 and 90 day mortality and an increased risk of requiring mechanical ventilation as compared to those of White ethnicity. The peak CRP and D-dimer levels in those of Black ethnicity were significantly higher than those of other ethnicities suggesting that these biological differences may accompany greater disease severity and increased risk of adverse outcomes.

*Implications of all the available evidence:* It is clear that ethnicity is a predictor of a positive SARS-CoV2 result, disease severity and mortality, regardless of age, sex, geographical location, deprivation, smoking status, BMI, co-morbidities and frailty. The association appears to be underpinned by a combination of factors including SES, pre-existing health conditions, biological risk factors such as D-dimers, environmental and structural determinants of health; but their relative contribution is unclear. Understanding these drivers is critical to designing interventions and refining clinical and Public Health policies. The evidence also emphasises the need for robust surveillance of ethnicity in health care research.

## Introduction

The novel *Severe Acute Respiratory Syndrome Coronavirus*-2 (SARS-CoV-2) which manifests as coronavirus disease 2019 (COVID-19) has led to a global pandemic(1). Older age, male sex, obesity and pre-existing health conditions such as diabetes and hypertension have all been identified as risk factors for poor outcomes(2-4). A disproportionate impact of disease severity and death on people from Black, Asian and minority ethnic (BAME) backgrounds has been reported, though not consistently. The UK Intensive Care National Audit and Research Centre (ICNARC) noted that whilst BAME groups only make up 14% of the UK population, they comprised 33% of COVID-19 patients on intensive care units(5). The degree of this excess risk also appears to differ across, and within, these heterogeneous ethnic groups. In the UK, recent analyses of data from the Office of National Statistics and NHS England described 2.5-4.3 fold greater COVID-19 mortality rates, compared to White groups, across a range of Black and South Asian ethnic groups(6). Whether this adverse association is driven by underlying co-morbid disease, socio-economic inequality, genetic factors or a complex interplay of them all is unclear(7). Current data are limited in either number of COVID-19 patients, ethnic diversity or event rates with limited adjustment for known risk factors and potential predictors(8-11). There is an urgent need for the detailed characterisation of ethnic differences in COVID-19 outcomes and associated risk factors, within diverse populations, to inform practice and policy. Identifying and responding to these ethnic inequalities will be key to mitigating the disproportionate impact of COVID-19 on BAME patients.

Barts Health NHS Trust is the largest NHS trust in the UK, comprising six hospitals; The Royal London Hospital, Newham General Hospital, Whipps Cross Hospital, Mile End Hospital (Non-acute), St Bartholomew’s Hospital and the London NHS Nightingale Hospital, a purposely built COVID-19 hospital. The hospitals serve the ethnically diverse and socially deprived communities of over 2.6 million people in east London including the London Borough of Newham which experienced 144.3 COVID-19 related deaths per 100,000 population(12), the highest mortality in the UK and Tower Hamlets which has the largest Bangladeshi population in England(13). This large, regional dataset afforded extensive analyses of COVID-19 patients of a higher acuity than other studies. We aimed to examine the demographic, socio-economic, behavioural, biochemical and clinical risk factors associated with outcomes within different ethnic groups of hospitalised COVID-19 patients, using multivariable survival analyses.

## Methods

### Study population

We considered all patients with confirmed SARS-CoV-2 infection and admitted to the five acute hospitals within Barts Health NHS Trust between 1^st^ January and 13^th^ May 2020. Diagnosis was made using one or more real-time RT-PCR. Those under 16 years were excluded. The first emergency admission encompassing the first positive SARS-CoV-2 test, or the first emergency admission within two weeks of positive outpatient testing was defined as the index admission, community diagnoses without an associated emergency hospital admission were excluded. Patients with unknown or undisclosed ethnicity status were collected for comparison but were not included in our primary ethnicity analysis.

### Data collection

Clinical and demographic data, blood results and coding data from current and prior clinical encounters, were collated from the Barts Health Cerner Millennium Electronic Medical Record (EMR) data warehouse and locally held ICNARC databases by members of the direct clinical care team. Mortality data was available to 20th May 2020.

### Definition of key variables

Ethnicity was defined using the NHS ethnic category codes and based on five high-level groups: White, Asian or Asian British, Black or Black British, Mixed and Other; to preserve statistical power the Mixed and Other categories were merged. Relative measures of socioeconomic deprivation were assessed using the English Indices of Deprivation 2020 by matching patient postcode to national index of multiple deprivation (IMD) quintiles using the Office of National Statistics Postcode Directory(14, 15). Baseline comorbid diseases and Hospital Frailty Risk Score (HFRS) were identified by mapping to ICD-10 coding(16). Body mass index (BMI) was calculated by height and weight measurements taken at or during the immediately preceding admission episode. Rockwood Clinical Frailty Scoring (RCFS) was assessed by the admitting medical team and recorded in the EMR(17). Secondary haemophagocytic lymphohistiocytosis (sHLH) risk score was calculated from peak values of blood results(18). Full definitions are detailed in supplementary materials.

### Outcomes

The primary outcome was 30-day mortality from time of index COVID-19 hospital admission. Secondary endpoints were 90-day mortality, ICU admission, ICU length of stay, duration of organ support on ICU, need for mechanical ventilation, hospital length of stay, and discharge destination if discharged alive from hospital.

### Statistical analyses

A prospective statistical analysis plan was developed(19). Baseline characteristics are presented as mean and standard deviation, median and interquartile range, or number and percentage, as appropriate. We compared proportions using Pearson’s Chi-square test or Fisher’s exact test and continuous variables using 2-sample t-test or Wilcoxon rank-sum test, as appropriate. Time-to-event analysis was undertaken with follow-up censored at 30 days, survivors with less than 30 days follow-up were censored at time of maximal follow-up. A Cox proportional hazards model was used to assess survival adjusted for age and sex. A further multivariable Cox model was developed to assess the effect of pre-defined risk factors described as associated with adverse outcomes in COVID-19: IMD quintile, smoking status, body mass index, diabetes, hypertension, and chronic kidney disease (CKD). The proportional-hazard assumption was assessed by inspection of scaled Schoenfeld residual plots and investigated by stratification(20). Logistic regression modelling of ethnicity on ICU treatment using mechanical ventilation was carried out. Effect measures are presented as hazard ratios (HR) or odds ratios (OR) with 95% confidence intervals (CI). All analyses were performed using R version 3.6.3 (R Core Team 2020).

### Sensitivity analyses

To assess the effect of including patients with incomplete clinical data, missing data for baseline risk variables included in the multivariable Cox model was imputed using Multivariate Imputation by Chained Equations(21). Additional multivariable models were also carried out using aggregate Charlson comorbidity index (CCI) as a measure of total comorbid disease burden, and HFRS or RCFS collected at hospital admission. Longer-term survival to 90 days was assessed using Cox proportional hazards modelling adjusted for age and sex.

## Results

A total of 1996 patients, aged 16 years and older, with a confirmed SARS-CoV-2 test result with an acute Barts Health admission on or before 13th May 2020 were included in this study [Figure S1]. The recruitment window encompassed the peak time period of COVID-19 diagnoses [Figure S2]. The majority of patients were classified as being in the two most deprived socio-economic quintiles in England. The ethnic distribution was White (n=703, 35·2%), Asian or Asian British (n=538, 27·0%), Black or Black British (n=340, 17·0%), Mixed and Other (n=156, 7·8%) and unknown or undisclosed (n=259, 13·0%).

### Population Characteristics

Baseline characteristics, interventions and outcomes across ethnic groups are shown in Table 1. Black and Asian ethnicity patients were significantly younger with a median age of 59 years (Asian) and 64 years (Black), compared to 73 years in the White group (p<0·001). Comorbidity data was available in 1700 (85.2%) of patients.

**Table 1.** Study population baseline characteristics stratified by ethnic group, n (%) unless otherwise stated. Total n=1996 unless otherwise stated. P values based on Chi-square (for categorical) or Kruskal-Wallis test (for continuous). SD: standard deviation, IQR: interquartile range, IMD: index of multiple deprivation, BMI: body mass index, TIA: transient ischaemic accident, HTN: hypertension, CKD: chronic kidney disease, sHLH: secondary haemophagocytic lymphohistiocytosis (without known underlying immunosuppression and bone marrow aspirate data), CRP: C-reactive protein, ICU: intensive care unit, RRT: renal replacement therapy.

|  | Stratified by ethnic group |  |  |  |  | p value |
| --- | --- | --- | --- | --- | --- | --- |
|  | Asian or Asian British | Black or Black British | Mixed and Other Ethnic Groups | White | Unknown and Undisclosed |  |
| n | 538 | 340 | 156 | 703 | 259 |  |
| **Age (years)** |  |  |  |  |  |  |
| Mean (SD) | 57.8 (18.5) | 64.2 (16.9) | 59.5 (17.2) | 69.4 (17.7) | 59.8 (16.5) | <0.001 |
| Median (IQR) | 59.0 (44.0-71.0) | 64.0 (53.0-79.0) | 59.0 (47.8-72.3) | 73.0 (58.0-84.0) | 61.0 (50.0-71.5) | <0.001 |
| **Male** | 332 (61.7) | 193 (56.8) | 103 (66.0) | 404 (57.5) | 178 (68.7) | 0.01 |
| **IMD quintile [n=1980]** |  |  |  |  |  | <0.001 |
| 1 (most deprived) | 139 (26.0) | 124 (36.7) | 50 (32.9) | 183 (26.2) | 66 (25.7) |  |
| 2 | 291 (54.5) | 165 (48.8) | 72 (47.4) | 269 (38.5) | 124 (48.2) |  |
| 3 | 49 (9.2) | 34 (10.1) | 20 (13.2) | 99 (14.2) | 44 (17.1) |  |
| 4 | 35 (6.6) | 9 (2.7) | 7 (4.6) | 86 (12.3) | 18 (7.0) |  |
| 5 (least deprived) | 20 (3.7) | 6 (1.8) | 3 (2.0) | 62 (8.9) | 5 (1.9) |  |
| **Smoking [n=1700]** | 30 (6.6) | 21 (7.1) | 10 (8.3) | 91 (14.8) | 21 (9.8) | <0.001 |
| **BMI [n=1248]** |  |  |  |  |  |  |
| Median (IQR) | 26.9 (24.1-31.1) | 28.2 (24.6-31.8) | 25.9 (23.1-29.0) | 26.3 (22.5-31.6) | 26.3 (22.5-30.8) | 0.04 |
| By category |  |  |  |  |  | 0.04 |
| <18.5 kg/m<sup>2</sup> | 9 (2.8) | 8 (3.6) | 1 (1.3) | 34 (6.9) | 11 (8.5) |  |
| 18.5 - <25 kg/m<sup>2</sup> | 101 (31.2) | 57 (25.3) | 31 (40.3) | 160 (32.5) | 43 (33.1) |  |
| 25 - <30 kg/m<sup>2</sup> | 114 (35.2) | 83 (36.9) | 27 (35.1) | 145 (29.5) | 40 (30.8) |  |
| 30 - <40 kg/m<sup>2</sup> | 87 (26.9) | 65 (28.9) | 17 (22.1) | 126 (25.6) | 28 (21.5) |  |
| ≥40 kg/m<sup>2</sup> | 13 (4.0) | 12 (5.3) | 1 (1.3) | 27 (5.5) | 8 (6.2) |  |
| **Co-morbidity using ICD-10 [n=1700]** |  |  |  |  |  |  |
| **Obesity** | 108 (23.6) | 82 (27.9) | 18 (14.9) | 161 (26.2) | 40 (18.7) | 0.01 |
| **Ischaemic heart disease** | 102 (22.3) | 62 (21.1) | 12 (9.9) | 149 (24.3) | 21 (9.8) | <0.001 |
| **Myocardial infarction** | 55 (12.0) | 23 (7.8) | 6 (5.0) | 83 (13.5) | 14 (6.5) | 0.002 |
| **Congestive heart failure** | 67 (14·7) | 54 (18·4) | 8 (6·6) | 114 (18·6) | 17 (7·9) | <0·001 |
| **Peripheral vascular disease** | 33 (7·2) | 35 (11·9) | 7 (5·8) | 67 (10·9) | 16 (7·5) | 0·06 |
| **Cerebral vascular accident or TIA** | 54 (11·8) | 54 (18·4) | 11 (9·1) | 157 (25·6) | 16 (7·5) | <0·001 |
| **Dementia** | 25 (5·5) | 27 (9·2) | 5 (4·1) | 103 (16·8) | 7 (3·3) | <0·001 |
| **Chronic obstructive pulmonary disease** | 119 (26·0) | 45 (15·3) | 18 (14·9) | 181 (29·5) | 34 (15·9) | <0·001 |
| **Diabetes** | 220 (48·1) | 157 (53·4) | 49 (40·5) | 179 (29·2) | 59 (27·6) | <0·001 |
| **HTN** | 261 (57·1) | 212 (72·1) | 64 (52·9) | 376 (61·2) | 96 (44·9) | <0·001 |
| **Moderate to severe CKD** | 92 (20·1) | 93 (31·6) | 16 (13·2) | 145 (23·6) | 17 (7·9) | <0·001 |
| **End-stage renal disease** | 39 (8·5) | 36 (12·2) | 7 (5·8) | 27 (4·4) | 4 (1·9) | <0·001 |
| **Liver disease** | 49 (9·1) | 24 (7·1) | 12 (7·7) | 58 (8·3) | 12 (4·6) | 0·25 |
| **Cancer** | 30 (6·6) | 26 (8·8) | 8 (6·6) | 68 (11·1) | 12 (5·6) | 0·04 |
| **Cancer with metastases** | 8 (1·8) | 5 (1·7) | 1 (0·8) | 22 (3·6) | 6 (2·8) | 0·18 |
| **Acquired immunodeficiency syndrome** | 0 (0·0) | 5 (1·7) | 0 (0·0) | 1 (0·2) | 0 (0·0) | 0·001 |
| **Charlson comorbidity index [n=1700]** |  |  |  |  |  | <0·001 |
| 0 | 131 (28·7) | 66 (22·4) | 42 (34·7) | 143 (23·3) | 91 (42·5) |  |
| 1-2 | 178 (38·9) | 100 (34·0) | 50 (41·3) | 203 (33·1) | 88 (41·1) |  |
| 3-4 | 70 (15·3) | 52 (17·7) | 16 (13·2) | 146 (23·8) | 20 (9·3) |  |
| ≥5 | 78 (17·1) | 76 (25·9) | 13 (10·7) | 122 (19·9) | 15 (7·0) |  |
| **Rockwood frailty Score [n=831]** |  |  |  |  |  | <0·001 |
| 1-2 (very fit, well) | 31 (15·9) | 6 (4·3) | 7 (14·9) | 36 (9·7) | 15 (18·8) |  |
| 3-4 (managing well, vulnerable) | 87 (44·6) | 51 (36·7) | 17 (36·2) | 118 (31·9) | 32 (40·0) |  |
| 5-6 (mildly to severely frail) | 65 (33·3) | 73 (52·5) | 18 (38·3) | 174 (47·0) | 29 (36·2) |  |
| 8-9 (very severely frail, terminally ill) | 12 (6·2) | 9 (6·5) | 5 (10·6) | 42 (11·4) | 4 (5·0) |  |
| **Hospital frailty Risk Score [n=1700]** |  |  |  |  |  | <0·001 |
| <5 (low risk) | 240 (52·5) | 123 (41·8) | 66 (54·5) | 197 (32·1) | 117 (54·7) |  |
| 5-15 (intermediate risk) | 132 (28·9) | 87 (29·6) | 38 (31·4) | 150 (24·4) | 73 (34·1) |  |
| ≥15 (high risk) | 85 (18·6) | 84 (28·6) | 17 (14·0) | 267 (43·5) | 24 (11·2) |  |
| **Baseline eGFR ml/min/1·72m<sup>2</sup> [n=1525]** |  |  |  |  |  |  |
| Median (IQR) | 72·8 (53·3-92·7) | 56·4 (36·2-80·2) | 75·6 (54·2-91·4) | 64·1 (46·2-82·0) | 78·2 (61·5-88·7) | <0·001 |
| eGFR <60 | 130 (29·6) | 135 (48·6) | 26 (26·0) | 239 (40·5) | 29 (24·6) | <0·001 |
| **Acute kidney injury first 7 days [n=1673]** | 98 (22·2) | 101 (34·7) | 32 (24·6) | 151 (24·4) | 48 (25·0) | 0·003 |
| *Blood results during admission* |  |  |  |  |  |  |
| **Highest creatinine $\mu\text{mol/L}$ [n=1691]** |  |  |  |  |  | <0.001 |
| Median (IQR) | 91.0 (72.0-157.0) | 119.0 (80.0-260.0) | 88.0 (71.8-120.3) | 98.0 (76.0-147.0) | 94.0 (75.0-132.0) |  |
| **Highest CRP [n=1761]** |  |  |  |  |  | <0.001 |
| Median (IQR) | 146.0 (72.0-287.8) | 181.5 (99.3-289.8) | 132.0 (66.0-226.0) | 136.0 (68.0-237.0) | 156.0 (75.5-272.5) |  |
| **Highest D-dimer mg/L [n=968]** |  |  |  |  |  | <0.001 |
| Median (IQR) | 1.0 (0.5-3.5) | 2.5 (0.9-10.3) | 1.1 (0.5-2.7) | 1.4 (0.6-3.4) | 1.5 (0.7-6.3) |  |
| **Highest sHLH score [n=1881]** |  |  |  |  |  |  |
| Mean (SD) | 31.1 (27.1) | 30.0 (27.9) | 27.6 (28.3) | 26.4 (24.8) | 32.1 (26.7) | 0.01 |
| *Intensive care unit (ICU)* |  |  |  |  |  |  |
| **ICU admission** | 108 (20.1) | 63 (18.5) | 28 (17.9) | 77 (11.0) | 85 (32.8) | <0.001 |
| **Days in hospital before ICU Mean (SD)** | 2.3 (5.2) | 2.9 (5.1) | 1.1 (1.8) | 2.3 (11.4) | 1.8 (4.2) | 0.75 |
| **ICU length of stay** |  |  |  |  |  |  |
| Median (IQR) | 8.0 (3.0-15.2) | 8.1 (3.5-14.1) | 8.5 (5.0-13.1) | 8.0 (3.9-12.0) | 10.0 (6.0-16.0) | 0.30 |
| **Mechanical ventilation within ICU admission** | 78 (72.2) | 50 (79.4) | 23 (82.1) | 59 (76.6) | 71 (83.5) | 0.40 |
| **RRT within ICU admission** | 28 (25.9) | 26 (41.3) | 7 (25.0) | 20 (26.0) | 18 (21.2) | 0.09 |
| *Days on organ support* |  |  |  |  |  |  |
| **Advanced respiratory Mean (SD)** | 11.0 (10.8) | 9.4 (8.8) | 8.2 (7.1) | 7.8 (7.8) | 10.3 (8.0) | 0.14 |
| **Total respiratory Mean (SD)** | 13.1 (10.4) | 11.9 (8.9) | 9.8 (7.0) | 9.6 (7.7) | 11.9 (7.6) | 0.08 |
| **Cardiovascular system Mean (SD)** | 13.4 (10.9) | 11.5 (8.6) | 9.9 (7.2) | 9.8 (8.3) | 11.8 (7.5) | 0.07 |
| **Renal Mean (SD)** | 2.4 (5.5) | 4.4 (6.6) | 2.1 (4.7) | 2.7 (5.7) | 1.5 (3.8) | 0.03 |
| **Total number of organ systems** |  |  |  |  |  | 0.15 |
| 0 | 1 (0.9) | 0 (0.0) | 0 (0.0) | 1 (1.3) | 1 (1.2) |  |
| 1 | 3 (2.8) | 4 (6.3) | 1 (3.6) | 5 (6.5) | 0 (0.0) |  |
| 2 | 76 (70.4) | 33 (52.4) | 20 (71.4) | 52 (67.5) | 66 (77.6) |  |
| 3 | 28 (25.9) | 26 (41.3) | 7 (25.0) | 19 (24.7) | 18 (21.2) |  |
| *Outcomes* |  |  |  |  |  |  |
| **Died** | 146 (27.1) | 101 (29.7) | 34 (21.8) | 230 (32.7) | 62 (23.9) | 0.01 |
| **Days to death** |  |  |  |  |  |  |
| Mean (SD) | 9.7 (10.0) | 9.1 (11.0) | 11.0 (9.8) | 12.9 (13.6) | 12.7 (10.0) | 0.02 |
| Median (IQR) | 6.0 (3.0-12.0) | 5.0 (3.0-11.0) | 10.5 (4.3-14.0) | 9.0 (4.0-16.0) | 10 (6.0-17.0) | <0.001 |
| **Died within 30 days** | 138 (25.7) | 97 (28.5) | 33 (21.2) | 210 (29.9) | 58 (22.4) | 0.05 |
| **Died within 90 days** | 146 (27.1) | 101 (29.7) | 34 (21.8) | 229 (32.6) | 62 (23.9) | 0.01 |
| **Still in hospital** | 7 (1.3) | 6 (1.8) | 3 (1.9) | 6 (0.9) | 5 (1.9) | 0.60 |
| **Hospital length of stay** |  |  |  |  |  |  |
| Median (IQR) | 5.0 (3.0-10.0) | 7.0 (4.0-12.0) | 5.0 (3.0-11.0) | 8.0 (4.0-15.0) | 8.0 (4.0-15.0) | <0.001 |
| **Discharged Hospital alive** | 402 (74.7) | 241 (70.9) | 122 (78.2) | 487 (69.3) | 200 (77.2) | 0.03 |
| **Discharge destination** |  |  |  |  |  | <0.001 |
| Care home or equivalent | 7 (1.8) | 5 (2.1) | 0 (0.0) | 40 (8.3) | 8 (4.0) |  |
| Health-related institution | 7 (1.8) | 10 (4.3) | 8 (6.7) | 23 (4.8) | 37 (18.7) |  |
| Usual place of residence | 373 (94.4) | 216 (91.9) | 110 (91.7) | 403 (83.8) | 152 (76.8) |  |
| Hospice or equivalent | 1 (0.3) | 0 (0.0) | 0 (0.0) | 2 (0.4) | 1 (0.5) |  |
| Temporary place of residence | 7 (1.8) | 4 (1.7) | 2 (1.7) | 13 (2.7) | 0 (0.0) |  |

Burden of comorbid disease varied between ethnic groups in prevalence, type and age-distribution. Overall distribution of COVID risk factors varied with age and ethnicity with diabetes and CKD more prevalent at an earlier age in Asian and Black patients and frailty and dementia more prevalent in older White patients [Figure 1].

**Figure 1.**
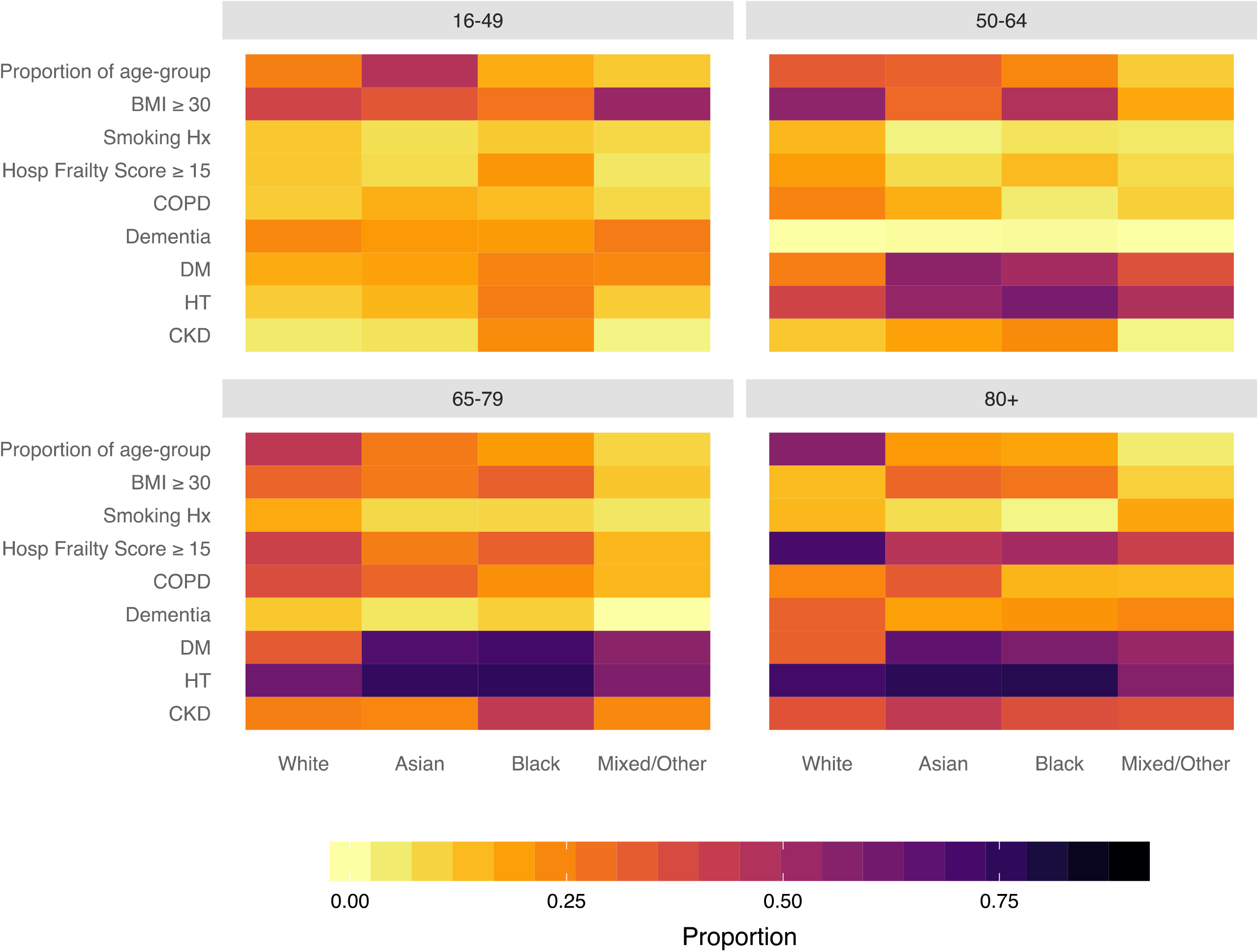
Heat map of prognostic factors in COVID-19 hospital admissions by age and ethnic background showing proportions within each ethnic group for each age group. Asian and Black patients differed from those of white background in the presence of risk factors and their age distribution however differences were also apparent between different Black and Minority Ethnic groups at different ages. Proportions are of those with data (see Table 1). BMI: body mass index, COPD: chronic obstructive pulmonary disease, DM: diabetes mellitus, HT: hypertension, CKD: chronic kidney disease.

**Figure 2.**
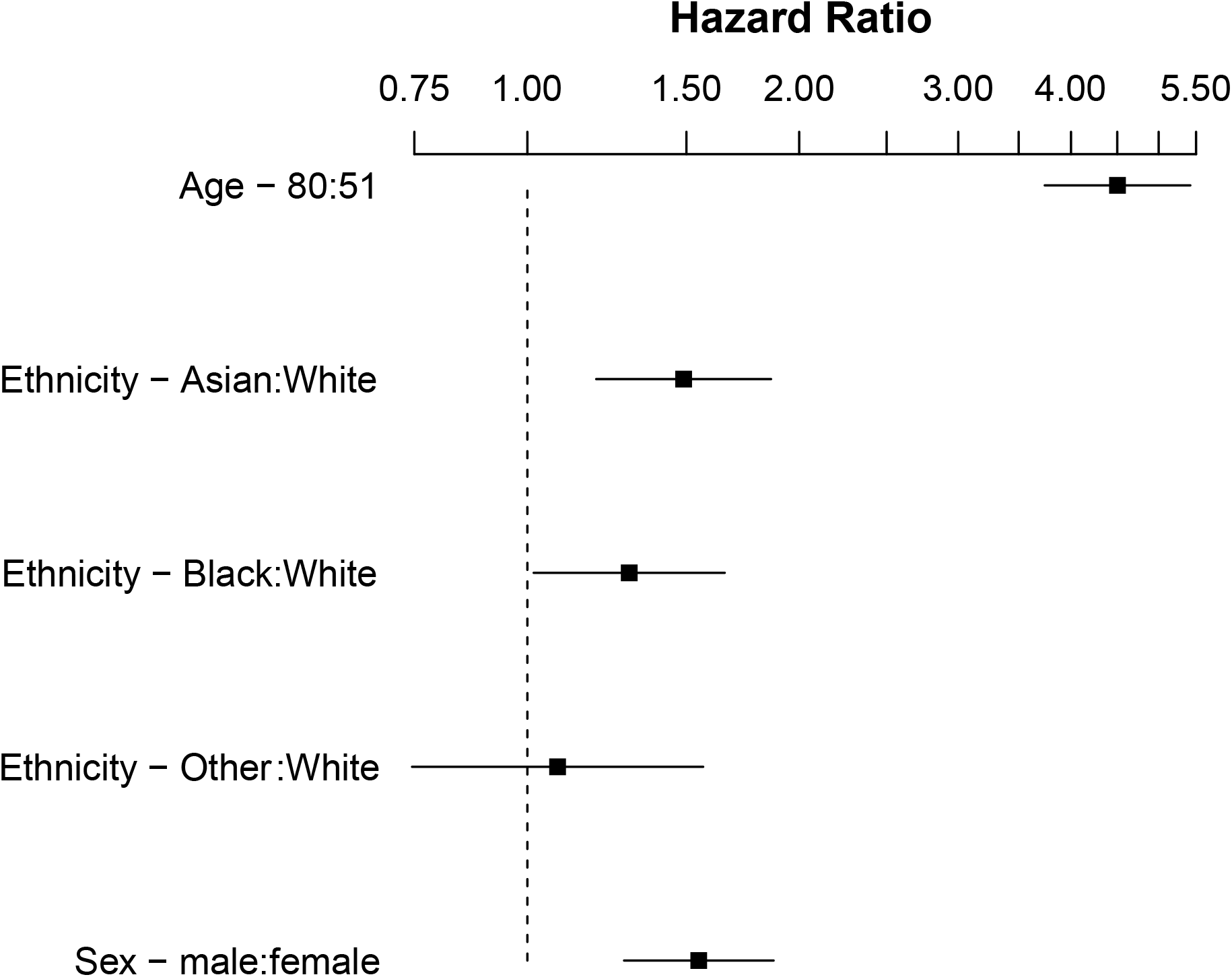
Forest plot showing log hazards ratios of mortality to 30 days comparing ethnic groups, age and sex corrected.

**Figure 3.**
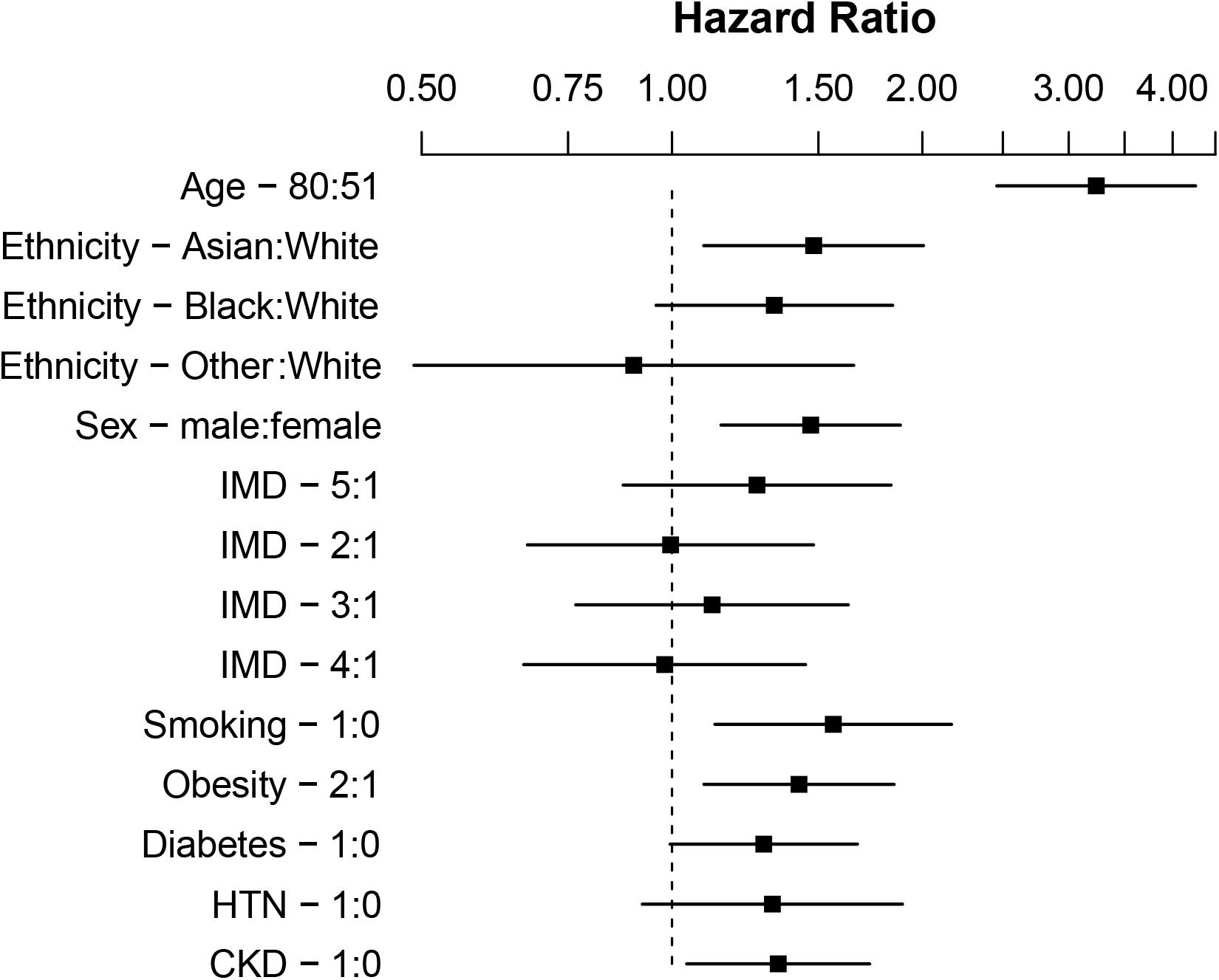
Forest plot showing log hazards ratios of mortality to 30 days comparing ethnic groups, age and sex corrected. Additional variables included index of multiple deprivation (IMD) quintile, smoking, BMI ≥30 kg/m^2^, diabetes, HTN: hypertension, CKD: chronic kidney disease.

**Figure 4.**
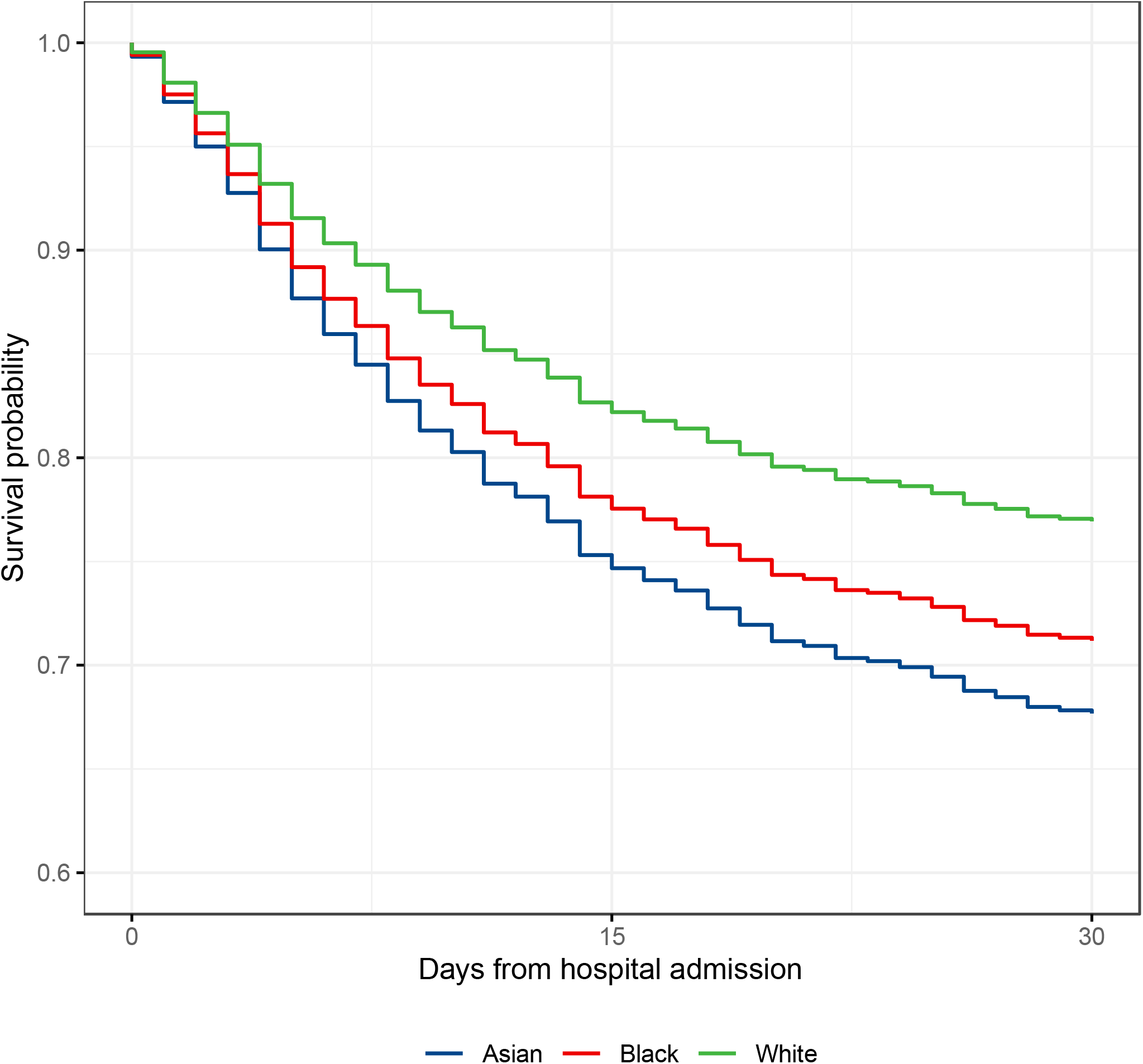
Survival curve to 30 days comparing predicted survival of Asian, Black, and White ethnic groups (Mixed and Other group omitted for clarity), in an age and sex adjusted Cox-hazard analysis. Survival curves adjusted to median age 65 years and male sex.

**Figure 5.**
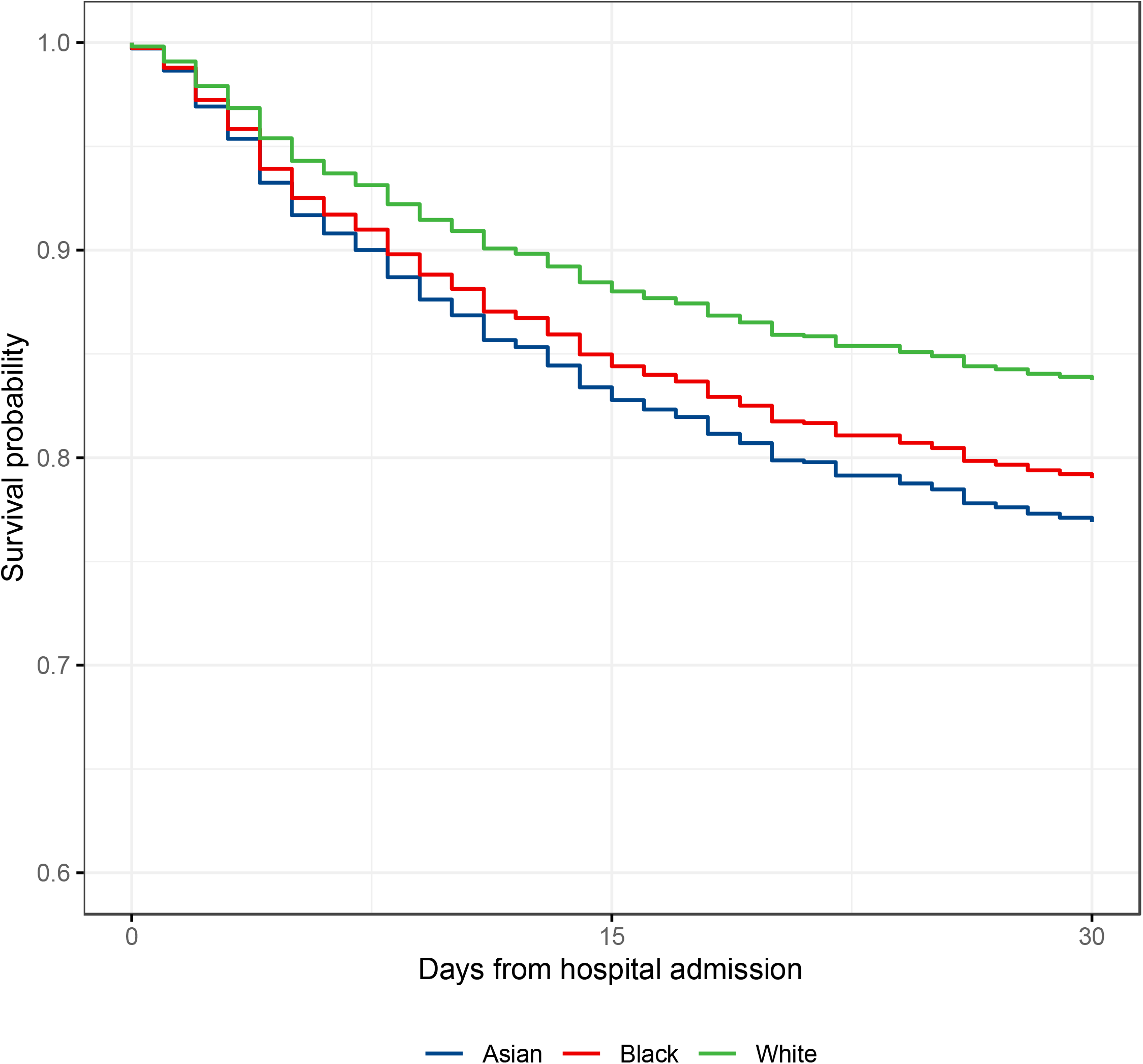
Survival curve to 30 days from multivariable analysis comparing Asian, Black, and White ethnic groups. Survival modelled for median age 65 years and male sex, index of multiple deprivation (IMD) least deprived quintile, no history of baseline risk factors defined as Non-smoking, BMI <30 kg/m^2^ and No diabetes, hypertension or chronic kidney disease. Statistically significant difference in survival between Asian group and White group persists after adjustment for age, sex, social deprivation and major COVID19 risk factors.

Around one in four patients developed early acute kidney injury (AKI) within seven days of hospital admission, rates of AKI were highest in the Black group (34·7%). Patients in the Black group had higher levels of inflammation measured using CRP (median 181·5 mg/L) and D-dimer (median 2·5 mg/L) compared to other ethnicities.

Total mortality to 20th May 2020 was 28·7% (n=573). Based on the raw data, a greater proportion of White patients died (32·7%) compared to Asian (21·1%) and Black (29·7%) patients. The majority of deaths (93·7%) occurred within 30 days of hospital admission.

### Age and sex adjusted 30-day mortality

We included 1737 Asian, Black and White patients in the primary outcome analysis. After adjustment for the between-group differences in age and sex, patients from Asian and Black ethnic groups were at significantly higher risk of death within 30 days compared to White patients (Asian ethnicity (HR 1·49, CI 1·19-1·86, p<0·001); Black patients (HR 1·30, CI 1·02-1·63, p=0·036). No association was observed in the smaller Mixed and Other Ethnicity group (HR1·08, CI 0·75-1·57, p=0·682) [Table 2]. There was some evidence of non-proportionality for the association between ethnicity and risk of death over time [Figure S14], consequently these HRs should be interpreted as a weighted average over the 30-day follow up period. To investigate change in risk over time we developed an ethnicity-stratified Cox-model, this supported the findings of the unstratified model, but suggested that Black ethnicity might be associated with a higher early rate of death [Figure S15].

**Table 2.** Association of ethnic group with mortality to 30 days using Cox proportional hazards modelling, age and sex corrected. Censored to 30 days follow up, observations 1737, events 478.

|  | n |  | Unadjusted |  |
| --- | --- | --- | --- | --- |
|  | Total | Events | Hazard ratio (95% CI) | p value |
| **Age** (25<sup>th</sup> vs 75<sup>th</sup> centile) | - | - | 4.50 (3.74-5.42) | <0.0001 |
| **Sex** (Male) | - | - | 1.55 (1.28-1.87) | <0.0001 |
| **Ethnic group** |  |  |  |  |
| Asian or Asian British | 521 | 134 | 1.49 (1.19-1.86) | <0.001 |
| Black or Black British | 331 | 94 | 1.30 (1.02-1.65) | 0.036 |
| Mixed and Other ethnic groups | 150 | 34 | 1.08 (0.75-1.57) | 0.682 |
| White | 674 | 206 | Reference | - |

### Multivariable survival modelling

After inclusion of IMD quintile, smoking history, BMI ≥30 kg/m^2^, diabetes, hypertension, and CKD in a multivariable survival analysis, the association with increased rate of death persisted in Asian patients (HR 1·48, CI 1·09-2·01, p=0·011; n=1006). In Black patients, the magnitude of the mortality trend was unchanged, however was outside the limits of standard statistical significance (HR 1·32, CI 0·96-1·84, p=0·090; n=1006), potentially due to the smaller sample size. In this model older age, male sex, smoking, BMI ≥30 kg/m2 and CKD were statistically associated with risk of death [Table 3] and there was no statistical evidence that ethnicity violated the proportional hazards assumption. The associations were broadly unchanged when the model was re-fitted after multiple imputation of missing values [Table S5].

**Table 3.**
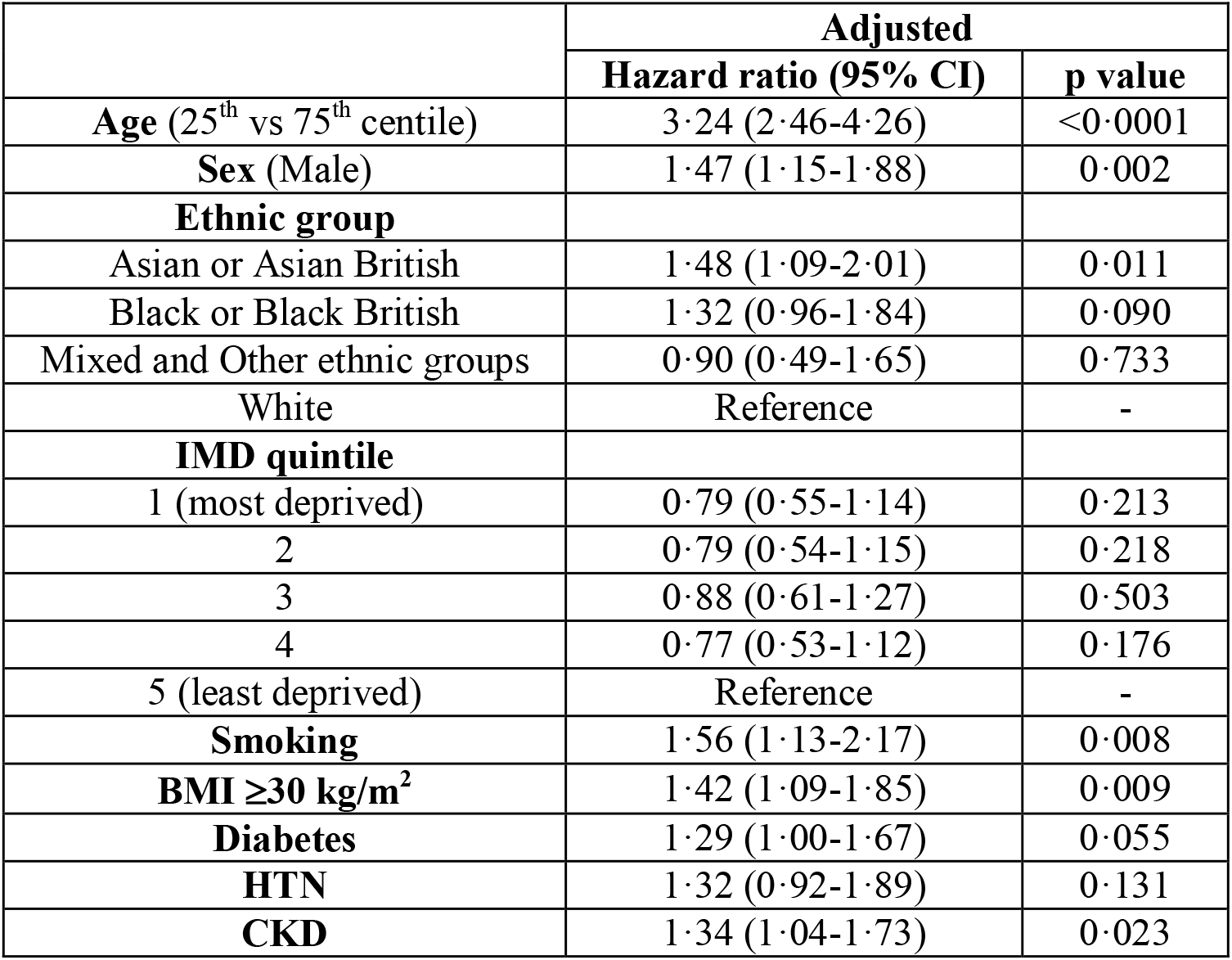
Multivariable analysis of mortality to 30 days using Cox proportional hazards modelling, age and sex corrected. Variables included IMD quintile, smoking, BMI ≥30 kg/m^2^, diabetes, HTN: hypertension, CKD: chronic kidney disease. Censored to 30 days follow up, observations 1006, events 281.

Sensitivity analyses for further multivariable survival models were developed to examine the influence of total comorbidity burden, as assessed by CCI [Table S6], and measures of frailty, the RCFS or HFRS [Tables S7, S8]. In all these analyses the association between Black and Asian ethnicity and 30-day mortality remained significant. Adjusting for RCFS raised the odds of 30-day mortality to a HR of 1·98 (CI 1·37-2·86; p<0.001) in Asian groups and to a HR of 1·67 (CI 1·14-2·45; p=0.009) in Black groups, with similar effect size in analysis adjusted for the HFRS [Tables S7, S8]. Asian ethnicity also continued to be associated with greater risks of death through to 90 days follow-up (HR 1·46, CI 1·18-1·81, p<0·001; n=1737) [Table S9].

### Critical Care related outcomes

In the White group, 11·0% of patients were admitted to ICU compared to 20·1% of the Asian group and 18·5% of the Black group (p<0.001). In those admitted to ICU, rates of mechanical ventilation requiring intubation did not differ significantly by ethnicity at 76·6% in the White group, 72·2% in the Asian group and 79·4% in the Black group. Similarly, while rates of ICU admission differed significantly, length of ICU stay did not with a median of 8-9 days in all ethnic groups. Across the entire hospitalised cohort Asian (OR 1·54, CI 1·06-2·23, p=0·023; n=1737) and Black (OR 1·80, CI 1·20-2·71, p=0·005; n=1737) ethnicities were associated with increased age and sex adjusted-risk of receiving invasive mechanical ventilation in ICU [Table S4]. There was a trend toward increased renal replacement therapy use in Black patients (41.3%) admitted to ICU compared to 20-25% across other ethnic groups (p=0.09).

## Discussion

We report on treatment and outcomes in COVID-19 patients hospitalised in East London throughout the peak of the UK pandemic, a population with the UK’s highest COVID-19 mortality. To our knowledge this is one of the largest UK hospital COVID-19 cohorts reported, and certainly the most diverse, with only 35.2% of 1996 patients identified as White ethnicity. We found those of Asian ethnicity to be at the highest risk of death within 30 days (HR 1·49, CI 1·19-1·86, p<0·001), a finding that persisted at 90 days. Risk of death in Black patients was also greater than those of White ethnicity (HR 1·30, CI 1·02-1·63, p=0·036). This disparity extended to need for ICU care with Asian and Black patients experiencing a 50-80% increased risk of receiving mechanical ventilation in ICU compared to White patients of a similar age.

### Findings in context

Our findings differ from previous reports in the UK and US in which Black ethnicity has been consistently associated with greater COVID-related mortality(6, 22). Preliminary analyses of the UK ICNARC report on COVID-19 in critical care highlighted Black ethnicity with the highest likelihood of being admitted to intensive care compared to a matched population (10.7% versus 6.5%)(23). Similarly, in a large UK primary care linked cohort, Black patients were also to found to be at highest risk of COVID-related death(9). In a US study, the composite relative risk of COVID-related death compared to White ethnicity was 3.57 in Black populations, and 1.88 for Latinos(22). Our findings suggest specific South Asian communities may have at least the same or higher risk in COVID-19 as those of Black background. This may reflect characteristics of the large South Asian, and specifically Bangladeshi, community in East London, poorly represented in other studies.

Older age has been significantly associated with increased COVID-19 mortality across a range of studies(2-4). In our cohort, patients from Asian and Black backgrounds were strikingly younger than White patients. However, despite the expected protective factor of younger age, when this was accounted for, those from Black and Asian backgrounds were more likely to die. The prevalence of co-morbid disease has been well described as a risk factor for COVID-19 disease and death(3, 4). We found different ethnic groups had differing age-distribution of baseline comorbidities such as hypertension, diabetes, chronic obstructive pulmonary disease and dementia. Despite accounting for these and other described predictors of poor outcomes, increased risk of death in Asian and Black populations was not attenuated, suggesting co-morbidities are not the sole drivers of ethnicity-associated risk.

Patients identified as frail have been predicted to have worse COVID-19 related health outcomes(24), and lower likelihood of benefiting from complex acute interventions, including critical care. In this study White patients, in addition to being notably older than other ethnicities, had higher degrees of frailty. Accounting for measures of frailty magnified the association seen between Asian and Black ethnicity and death. This suggests that whilst in White patients COVID-19 related death may have occurred in already frail and functionally vulnerable patients, in both Asian and Black patients, COVID-19 related deaths are likely to be occurring prematurely, in younger, fitter individuals with less functional vulnerability.

In our cohort, all ethnic groups experienced high levels of deprivation, however, worse deprivation was not associated with higher likelihood of mortality suggesting ethnicity may affect outcomes independent of purely geographical and socio-economic factors(25).

We found evidence for worse disease severity in Black and Asian groups as evidenced by higher rates of ICU admission and higher rates of AKI, and high levels of D-dimers and CRP in Black patients. High CRP and D-dimer levels have been identified as important inflammatory markers which strongly correlate with COVID-19 disease severity and prognosis(26). Our data suggest potential biological differences in host-response to COVID-19 may occur between ethnicities, however, causative associations in determining COVID-19-related mortality have not been demonstrated.

Finally, although COVID-19 has cast the effects of ethnic inequalities on health outcomes into sharp focus, these inequalities are not new. Health inequalities within and between ethnic minority groups are widely documented and the effects of structural racism are transmitted across generations(27). The risk factors already discussed such co-morbidity and obesity are speculated to intersect and be inextricably linked with wider social determinants such as poor living conditions, key worker roles and language barriers which impede the adoption of preventative measures(25, 28, 29). Some researchers have postulated that ethnicity may be associated with decreased symptom recognition and poor health literacy resulting in delayed presentation for care-something that could contribute to the greater illness-severity we observed(30).

### Strengths and Limitations

We believe this study is both one of the largest and most detailed of studies exploring COVID-19 outcomes in BAME populations so far reported. In contrast to previous studies examining ethnicity and COVID-19 outcomes we were able to address the contributions of socio-economic deprivation, comorbid disease, pre-morbid function, lifestyle and demographic factors to ethnic disparities in COVID-19 outcomes, including ICU interventions. Our analysis was strengthened by the inclusion of measures of frailty which is a critical determinant of outcomes in acute disease as well as a potential driver of clinician decision-making. It should be acknowledged, however, that frailty has social and biological dimensions and measures have not been extensively validated in BAME groups.

Importantly, this study was conducted in a single region where COVID-19 has had significant impact and thus is not confounded by differences in incidence of COVID-19 disease across the UK, regional concentration of minority ethnic groups and regional differences in the time-course of the epidemic. In addition, we employed a pre-specified statistical analysis plan and performed multiple sensitivity analyses to test the robustness of our findings.

Limitations in our analyses must also be considered. Importantly, SARS-CoV-2 testing has an appreciable false negative rate and suspected, but not proven, cases are an important group. Nevertheless, given that clinical suspicion varied both between cases and across the time-course of the epidemic with coding of suspected cases being inconsistent, in line with the vast majority of published COVID-19 analyses, we only included proven COVID-19 cases. Testing was available for all hospitalised patients with suspected COVID-19 disease, so availability of testing was not a bias. However, suspected diagnoses should be considered in future studies, particularly those occurring outside of hospitals, where not all clinical diagnoses may have been tested.

Similar to many hospital datasets there were missing data for a proportion of co-variates(8, 9), however 85% of patients had coding data for assessment of comorbidity and 63% measured height and weight data, providing a large sample with detailed data for analysis. We also imputed missing data and performed sensitivity analyses on our multivariable comorbidity models. This reinforced the observed ethnic differences, providing further confidence that our findings were not affected by missing data.

Like many datasets, our ethnic categorisations were aggregated and did not reflect the vast heterogeneity within ethnic categories (such as Bangladeshi, Pakistani, Black African or Black Caribbean). Indeed, the descriptive term “ BAME” itself is particularly crude and we recognise its limitation. Despite its size, our study lacked the power to assess a more detailed ethnicity breakdown. In addition, our observations in those of Asian ethnicity are likely skewed by our large Bangladeshi community, which has specific socio-economic and healthcare inequalities. It is therefore important that, suitably powered, analyses are conducted to expose differences between sub-ethnic categories. Similarly, whilst we have explored socio-economic factors, our analysis does not allow us to contextualise a number of potential socio-spatial factors including household composition, environmental factors and occupation. These should be considered in future research.

## Conclusion

In this analysis of a large, ethnically diverse and socio-economically challenged cohort, hospitalised patients of Asian and Black background with COVID-19 were at increased risk of premature death, independent of frailty, co-morbidities and social deprivation. Failure to robustly respond to the ethnic disparities so conspicuously unmasked during the COVID-19 pandemic can only further entrench and inflict them on future generations.

## Data Availability

The statistical analysis plan can be accessed online. The authors will be happy to consider additional analyses of the anonymised dataset on request. The need for stringent measures to prevent re-identification of individuals within a discrete geographical location and limited time-period however preclude sharing of patient level dataset in a GDPR compliant form.

## Contributions

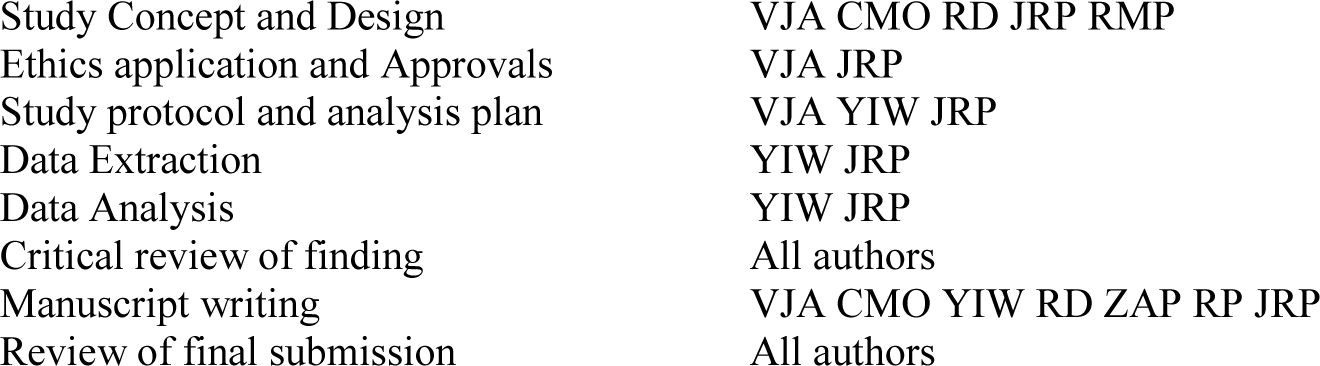

## Acknowledgments

We would like to acknowledge the assistance of the ICU clinical leads and ICU audit teams at the individual sites for access to local audit data: Gail Marshall (RLH and NGH), Susan Kolakottu Thomas (NUH), Jonathan Barry (SBH), John Sant (WXH). JP would like to thank Dr Bhavi Trivedi, Barts Health Clinical Analytics Officer for her past and current advice on data extraction from Trust EMR systems.

More widely we would like to acknowledge the courage and commitment of all staff members of Barts Health NHS Trust during the COVID-19 epidemic and to extent our condolences to all affected by death or serious illness related to COVID-19. The strength and character of the East London community from every background has once again been demonstrated.

